# Of masks and methylene blue - the use of methylene blue photochemical treatment to decontaminate surgical masks contaminated with a tenacious small non-enveloped norovirus

**DOI:** 10.1101/2021.11.04.21265909

**Authors:** Constance Wielick, Allyson Fries, Lorène Dams, Ravo M. Razafimahefa, Belinda Heyne, Brian H. Harcourt, Thomas S. Lendvay, Jean François Willaert, Simon de Jaeger, Eric Haubruge, Etienne Thiry, Louisa F. Ludwig-Begall

## Abstract

**Background:** In the context of the SARS-CoV-2 pandemic, reuse of personal protective equipment, specifically that of medical face coverings, has been recommended. The reuse of these typically single-use only items necessitates procedures to inactivate contaminating human respiratory and gastrointestinal pathogens. We previously demonstrated decontamination of surgical masks and respirators contaminated with infectious SARS-CoV-2 and various animal coronaviruses via low concentration- and short exposure methylene blue photochemical treatment (10 µM methylene blue, 30 minutes of 12,500-lux red light or 50,000 lux white light exposure).

**Methods:** Here, we describe the adaptation of this protocol to the decontamination of a more resistant, non-enveloped gastrointestinal virus and demonstrate efficient photodynamic inactivation of murine norovirus, a human norovirus surrogate.

**Results:** Methylene blue photochemical treatment (100 µM methylene blue, 30 minutes of 12,500-lux red light exposure) of murine norovirus-contaminated masks reduced infectious viral titres by over four orders of magnitude on surgical mask surfaces.

**Discussion and Conclusions:** Inactivation of a norovirus, the most difficult to inactivate of the respiratory and gastrointestinal human viruses, can predict the inactivation of any less resistant viral mask contaminant. The protocol developed here thus solidifies the position of methylene blue photochemical decontamination as an important tool in the package of practical pandemic preparedness.

## INTRODUCTION

In the context of the ongoing severe acute respiratory syndrome coronavirus 2 (SARS-CoV-2) pandemic, the supply of personal protective equipment (PPE) remains under strain. In a turn from the prevailing culture of throwaway living towards a sustainable circular economy within the healthcare industry [1,2], re-use of typically single-use only face coverings such as surgical face masks (SMs) and filtering facepiece respirators (FFRs) has been recommended [3,4]. Prior decontamination is paramount to safe PPE re-use and must guarantee the complete inactivation of SARS-CoV-2 as well as that of other contaminating respiratory or gastrointestinal human pathogens [5]. This is relevant in current circumstances (contamination with pathogens other than SARS-CoV-2 might easily occur, particularly in the context of widespread mask use and inexpert donning and doffing), but also plays a significant role in positioning the world for future pandemics [6].

Human respiratory pathogens include other enveloped corona-, pneumo-, metapneumo-, paramyxo-, and orthomyxoviruses as well as non-enveloped coxsackie- and rhinoviruses; gastrointestinal pathogens include boca-, astro-, picorna-, rota- and noroviruses (all non-enveloped) [7]. While enveloped viruses, surrounded by an outer lipid layer, are susceptible to inactivating treatments, non-enveloped viruses are known to be significantly more resistant. Amongst them, the small, non-enveloped human norovirus (genus *Norovirus*, family *Caliciviridae*), recognised as the major global cause of viral gastroenteritis [8], is notorious for its tenacity in the face of decontamination and as such may be considered the gold standard for validating viral inactivation [9,10]. The genetically and structurally similar murine norovirus (MNV), which replicates efficiently *in vitro*, has been identified as an appropriate surrogate for modelling human norovirus inactivation [11].

We previously demonstrated efficient inactivation of both a SARS-CoV-2 surrogate and an infectious animal norovirus via hydrogen peroxide-, ultraviolet germicidal irradiation-, and dry heat decontamination [12–14]. Particularly, the former two technologies, while easily deployable and extremely useful in high-resource settings, are not equitable as they remain less available in low-resource settings; accessible alternative decontamination methods are thus necessary to mitigate PPE shortages in restricted surroundings.

To address this issue, the Development and Methods for N95 Respirators and Mask Decontamination (DeMaND) study recently established a low-cost methylene blue (MB) photochemical treatment for the efficient decontamination of SMs and FFRs contaminated with infectious SARS-CoV-2 or surrogate animal coronaviruses [15]. Photosensitive MB dye, FDA-approved as an oxidation-reduction agent for the treatment of acquired methaemoglobinemia, has a long-standing history of use in pathogen inactivation both in the context of plasma treatment [16–18] and maxillary sinusitis therapy [19]. For its application to photochemical PPE decontamination, contaminated materials were coated with MB and subsequently exposed to a visible light source triggering the generation of virucidal singlet oxygen. A 10 µM MB concentration and a 30-minute exposure to 12,500 lux (10.474 W/m^2^) of red light or 50,000 lux (39 W/m^2^) of white light reduced titres of SARS-CoV-2 and two surrogate viruses by more than three orders of magnitude on all tested materials [15].

In the present investigation into decontamination of virus-inoculated SMs, we demonstrate inactivation of a highly resistant, small non-enveloped norovirus via MB photochemical treatment. Decontamination via a protocol adapted from the DeMaND study and involving the spray-coating of SMs with a 100 µM MB solution followed by 30 minutes of 12,500-lux red light exposure, robustly reduced infectious norovirus titres by over four orders of magnitude on SM coupons, this in excess of the three orders of magnitude reduction outlined in the current FDA policy regarding face masks and respirators [5]. This study serves to future-proof MB photochemical treatment since inactivation of a norovirus, the most resistant of the respiratory and gastrointestinal human viruses, can predict the inactivation of any less resistant viral mask contaminant. The protocol developed here thus solidifies the position of MB decontamination as an important tool in the package of practical pandemic preparedness.

## METHODS

### Viruses and cells

Murine macrophage cell line RAW264.7 (ATCC TIB-71) was maintained in Dulbecco’s modified Eagle’s medium (Invitrogen) complemented with 10% heat inactivated foetal calf serum (FCS) (BioWhittaker), 2% of an association of penicillin (5000 SI units/mL) and streptomycin (5 mg/mL) (PS, Invitrogen) and 1% 1 M HEPES buffer (pH 7.6) (Invitrogen) (DMEMc) at 37 °C with 5% CO_2_. Stocks of MNV isolate MNV-1.CW1 were produced by infection of RAW264.7 cells at a multiplicity of infection of 0.05. Two days post-infection, cells and supernatant were harvested and clarified by centrifugation for 20 minutes at 1000 x g after three freeze/thaw cycles (– 80°C/37°C). Titres were determined via TCID_50_ method; RAW 264.7 cells were seeded in 96-well plates, infected with ten-fold serial dilutions of MNV, incubated for three days at 37 °C with 5% CO_2_, and finally stained with 0.2% crystal violet for 30 minutes. Titres, expressed as TCID_50_/mL, were calculated according to the Reed and Muench transformation [22]. A virus stock with a titre of 7.06 log_10_ TCID_50_/mL was used in subsequent steps.

The continuous swine testicle (ST) cell-line, grown from testicular foetal swine tissues as described by McClurkin and Norman (1966) [20], was maintained in MEM (GIBCO), supplemented with 5% foetal calf serum (FCS) (Sigma), 1% sodium pyruvate 100x (GIBCO), and antibiotics (100U/mL penicillin, 0.1mg/mL streptomycin and 0.05 mg/mL gentamycin). Porcine respiratory coronavirus (PRCV) strain 91V44 [21] was passaged three times on confluent ST monolayers. Titres were determined via the tissue culture infective dose (TCID_50_) method; ST cells were seeded in 96-well plates and infected with ten-fold serial dilutions of PRCV and incubated for four days at 37 °C with 5% CO_2_. Four days after inoculation, monolayers were analysed for the presence of cytopathic effect by light microscopy. Titres, expressed as TCID_50_/mL, were calculated according to the Reed and Muench transformation [22]. A PRCV stock with a titre of 7.80 log_10_ TCID_50_/mL was used in subsequent steps.

### Surgical masks

Type IIR-Class I three-layer medical masks manufactured by Zarena AD, Bulgaria (LOT 0420; REF FMN99) were utilised in all assays. All masks, verified to be from the same manufacturing lot, were supplied by the Department of the Hospital Pharmacy, University Hospital Centre of Liege (Sart-Tilman).

### Light box

The light box designed by the Terra Research Centre and previously used for MB activation and PRCV decontamination in the context of the DeMaND study [15] contains six 180 W horticultural LED lamps (Roleadro Culture Indoor IP65 LED Horticultural T5 Grow), which together emit 12,500 LUX (or 10.474 W/m²) at 660 nm wavelength (luminescence verified using a light metre (DeltaOHM, Model HD2102.2)). The box interior is cooled by a ventilator equilibrating temperatures within the box (thus eliminating possible temperature effects on viral titres). Exposure to the red light within the light box (LB) is henceforth referred to as “LB exposure”. The design specifications of the light box have been made available in open source.

### Validation of previously established methylene blue photochemical decontamination using a porcine coronavirus and evaluation of its efficacy in inactivating a murine norovirus

To validate previously published MB photochemical treatment conditions (10 µM MB and 30-minutes of LB exposure) and to set a “baseline” for further assays involving MNV inocula, the previously established DeMaND protocol was investigated on SMs experimentally inoculated with PRCV. The same protocol was then tested for decontamination of MNV-inoculated SMs. The general workflow followed previously described protocols for PRCV or MNV [12–14] inoculation, MB photochemical treatment [15], and virus elution from SMs [12–14] and is also described in more detail below. Briefly, following injection of 100 µl undiluted PRCV or MNV suspension (7.80 and 7.06 log_10_ TCID_50_/mL, respectively; verified via back-titration) under the first outer layer of designated SM coupons, inoculated SMs were comprehensively sprayed with 7 – 8 mL of a 10 µM MB solution (Sigma-Aldrich (M9140); solution prepared in deionised water; total dose of 0.024 mg MB per SM), allowed to dry, and were then either kept in the dark or submitted to LB exposure for a duration of 30 minutes. Upon completion of the decontamination protocol, PRCV or MNV was eluted from the excised coupons and titres of recovered virus were determined via TCID_50_ assay in ST (PRCV) or RAW264.6 (MNV) cells. Methylene blue only controls (cytotoxicity test) and virus only controls (positive control) accompanied these assays.

### Establishment of concentration- and time-dependent virucidal kinetics of methylene blue photochemical treatment on murine norovirus

#### Virucidal methylene blue and light kinetics - microplate assay

To establish optimal treatment conditions and determine potential combined cytotoxic effects of higher concentrations of MB and light (MBL) on RAW264.7 cells, concentration- and LB exposure time-dependent virucidal MBL kinetics were investigated in a set of preliminary microplate-based assays. Ten-fold MB dilutions in deionised water (at final concentrations of 10 µM, 100 µM, and 1000 µM) in a total volume of 500 µL DMEMc (additionally supplemented with 0.1% beta-mercaptoethanol) were added per well of a 48-well plate containing 10 µL MNV (7.80 log_10_ TCID_50_/mL) (technical triplicates were performed utilising separate 48-well plates). The plates were then either kept in the dark (0 minute LB exposure) or were subjected to LB exposure for 30 minutes, 60 minutes, 90 minutes, or 120 minutes. In a second step, 100 µM and 1000 µM MB concentrations were tested against high-titre MNV (8.55 log_10_ TCID_50_/mL) with LB exposure times of 1 hour, 2 hours, 3 hours, and 4 hours. Methylene blue only controls (cytotoxicity test) and virus only controls (positive control) accompanied each assay. Titres of infectious MNV recovered from individual wells were determined via TCID_50_ assay. Back titrations of inoculum stocks were performed in parallel to each series of decontamination experiments.

#### Virucidal methylene blue kinetics - surgical mask assay

To establish optimal applied treatment conditions, LB exposure time- and concentration-dependent virucidal kinetics of MB were investigated on SMs experimentally inoculated with MNV.

##### Murine norovirus inoculation onto surgical masks

The workflow followed previously described protocols for SM inoculation with MNV [12,14]. Per SM, 100 µl of undiluted viral suspension were injected under the first outer layer at the centre of each of three square coupons (34 mm x 34 mm). The SMs were allowed to dry for 20 minutes at room temperature before decontamination via MB photochemical treatment.

##### Methylene blue photochemical treatment of surgical masks

Inoculated SMs were placed horizontally and were sprayed with a total volume of 7-8 mL of either a 100 µM or 1000 µM MB solution (the final MB volume was determined based on six repetitive sprays into a graduated cylinder). In total, each SM was sprayed four times on the outer side and twice on the inner side (facing the wearer). Surgical masks were allowed to dry (absorption of the MB solution) for 30 minutes in a dark box and were then either kept in the dark (0-minute LB exposure) or were subjected to LB exposure for 120 minutes, 180 minutes, or 240 minutes. Methylene blue only controls (cytotoxicity test) and virus only controls (positive control) again accompanied each assay.

##### Murine norovirus elution from surgical masks

Upon completion of the decontamination protocol, downstream coupon excision and virus elution followed previously described protocols [12,13]. Briefly, MNV was eluted from three excised coupons per SM into 4 mL DMEMc via a 1-minute vortex at maximum speed (2500 rounds per minute; VWR VX-2500 Multi-Tube Vortexer). Titres of infectious MNV recovered from individual coupons were determined via TCID_50_ assay. Back titrations of inoculum stocks were performed in parallel to each series of decontamination experiments.

#### Validation of a virucidal photochemical methylene blue treatment to decontaminate murine norovirus – inoculated surgical masks

To verify a treatment protocol wherein MNV-inoculated SMs (7.5 log_10_ TCID_50_/mL) were sprayed with 100 µM MB and subjected to LB exposure for 30 minutes, four biological and technical repeats were performed on four different days. Methylene blue only controls (cytotoxicity test) and virus only controls (positive control) accompanied these assays.

#### Testing shorter light box exposure (15 minutes) for photochemical methylene blue decontamination of murine norovirus – inoculated surgical masks

In an additional step, 100 µM MB concentrations were tested against MNV-inoculated SMs (7.30 log_10_ TCID_50_/mL) in conjunction with a shorter LB exposure time of 15 minutes (biological and technical triplicates). Methylene blue only controls (cytotoxicity test) and virus only controls (positive control) again accompanied these assays.

## Data analysis

Differences in infectious viral titres were computed and all graphs created using GraphPad Prism 7 (Graph-Pad Software). Statistical analyses of differences in infectious viral titres were performed using GraphPad Prism 7 (Graph-Pad Software) and P-values were computed by using a two-sided independent sample t-test, where ****P<0.0001, ***P<0.001, **P<0.01, *P<0.05, and ns is P≥0.05.

## RESULTS

### Methylene blue photochemical treatment of surgical masks following previously established protocols for coronavirus inactivation reduces porcine respiratory coronavirus titres by over five orders of magnitude but does not inactivate murine norovirus

Photochemical treatment involving application of a 10μM MB solution and a 30-minute LB exposure reduced infectious PRCV titres to below the assay detection limit of 0.8 log_10_ TCID_50_/mL on SM coupons (5.33 (±0.25) log_10_ TCID_50_/mL reduction. Following the same treatment protocol, MNV titres dropped from 5.78 (±0.33) log_10_ TCID_50_/mL to 5.19 (±0.16) log_10_ TCID_50_/mL, resulting in a total reduction of 0.53 (±0.34) TCID_50_/mL (Figure 1).

**Figure 1.**
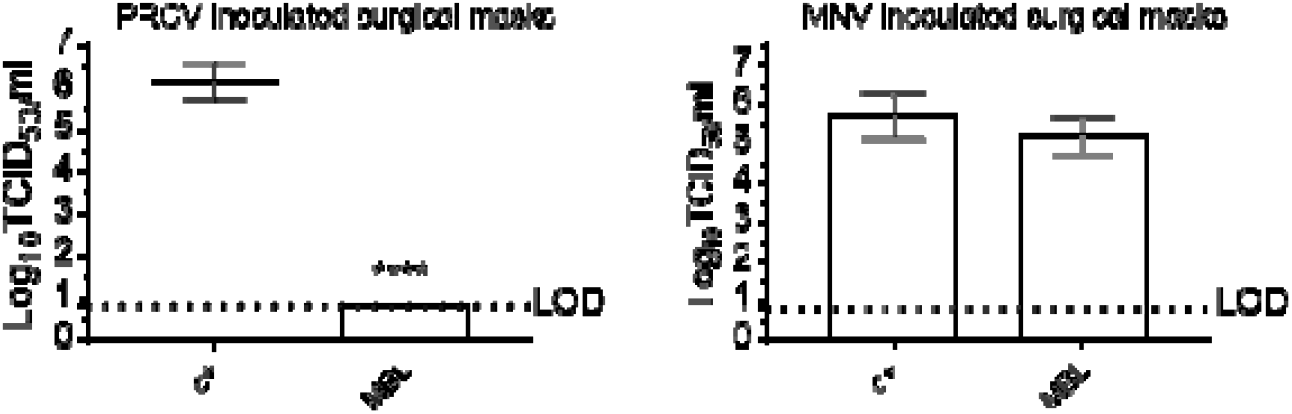
Validation of the previously established methylene blue (MB) photochemical treatment protocol using an enveloped animal coronavirus (left panel) and evaluation of its efficacy in inactivating a small non-enveloped norovirus (right panel). Porcine respiratory coronavirus (PRCV) - or murine norovirus (MNV) - inoculated surgical mask coupons remained untreated (c+) or were treated with a 10 µM MB solution and exposed to a 12,500-lux red light source (light box) for 30 minutes (MBL). The infectivity of PRCV recovered from surgical mask coupons (n=3) was analysed in swine testicular cells. The infectivity of MNV recovered from mask coupons (n=9) was analysed in RAW 264.7 cells. Values for positive controls (MB-untreated, but light box-exposed) ranged between 5.80 and 6.63 log_10_ TCID_50_/mL (PRCV) 5.05 and 6.05 log_10_TCID_50_/mL (MNV). The cell culture limit of detection (LOD) was 0.80 log_10_ TCID_50_/mL for all analyses. Means log_10_ TCID_50_/mL and standard deviations are represented. P-values were computed by using a two-sided independent sample t-test (where ****P<0.0001).

### Murine norovirus inactivation in microplates via methylene blue photochemical treatment is concentration- and time-dependent

To establish optimal MNV treatment conditions, a preliminary microplate-based assay examined concentration- and LB exposure time-dependent virucidal kinetics of MB photochemical treatment. Again, treatment with 10 µM MB concentrations and 30 minutes of LB exposure (treatment conditions previously utilised to successfully inactivate SARS-CoV-2 and its surrogates by more than three orders of magnitude [23]) reduced viral titres by less than one order of magnitude (0.73 (±0.53) log_10_ TCID_50_/mL). Protocols combining the same LB exposure time with 100 µM and 1000 µM MB concentrations both reduced MNV titres by 0.93 (± 0.35) log_10_ TCID_50_/mL. With increasing exposure times, viral titres successively dropped, reaching absolute titre reductions of 1.73 (± 0.53), 2.68 (± 0.35), and 2.50 (± 0.00) log_10_ TCID_50_/mL after 120 minutes (2 hours) of LB exposure in conjunction with 10 µM, 100 µM, and 1000 µM MB concentrations, respectively. Ten-fold increases in MB concentrations coincided with higher MB cytotoxicity and correspondingly elevated assay detection limits (LODs) from 0.8 to 1.8, to 2.8 log_10_ TCID_50_/mL (Figure 2).

**Figure 2.**
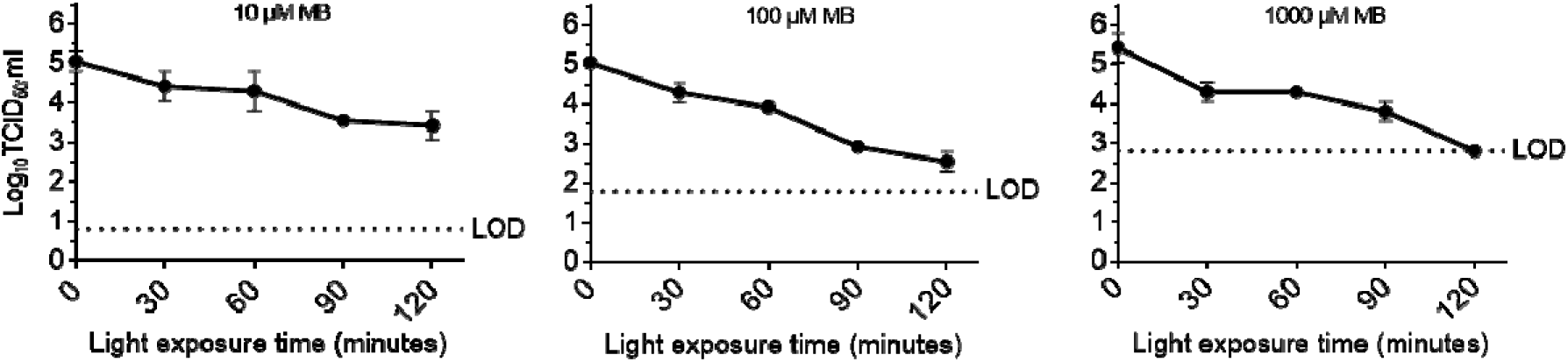
Evaluation of concentration- and time-dependent virucidal kinetics of methylene blue (MB) photochemical treatment on murine norovirus (MNV) *in vitro*. The infectivity of MNV recovered from microplate wells containing medium and MB at different concentrations and exposed to a 12,500-lux red light source (light box) was analysed in RAW 264.7 cells. The cell culture limit of detection (LOD) was 0.80, 1.8, and 2.8 log_10_ TCID_50_/mL for 10 µM MB, 100 µM MB, and 1000 µM MB concentrations, respectively. All assays were performed as technical duplicates (n=2). Means log_10_ TCID_50_/mL and standard deviations are represented. Values for positive controls (MB-untreated, but light box-exposed MNV) ranged between 4.80 and 5.30 log_10_ TCID_50_/mL.

In a second microplate-based assay, 100 µM and 1000 µM concentrations were tested against high-titre MNV in conjunction with longer LB exposure times of 1 hour, 2 hours, 3 hours, and 4 hours. Following 2 hours of LB exposure, virus titre reductions mirrored those observed in the first assay at this time point (2.69 (± 1.66) and 2.85 (± 1.58) log_10_ TCID_50_/mL after treatment with 100 µM and 1000 µM MB solutions). Following treatment with a 100 µM MB solution and 3 hours of LB exposure, viral titres dropped by 4.80 (± 1.01) log_10_ TCID_50_/mL; treatment with 100 µM MB and 4 hours of LB exposure as well as a 1000 µM MB followed by 3 and 4 hours of LB exposure lowered viral titres to below the LODs of each of the respective assays, thus yielding minimal titre reductions of 3.89 (± 0.90), 3.22 (± 1.45), and 2.89 (± 1.40) log_10_ TCID_50_/mL, respectively (Figure 3).

**Figure 3.**
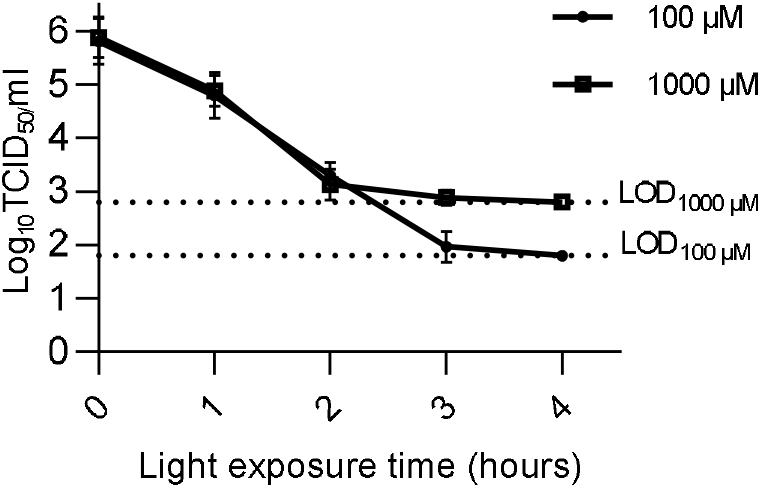
Evaluation of concentration- and longer exposure time-dependent virucidal kinetics of methylene blue (MB) photochemical treatment on murine norovirus (MNV) *in vitro*. The infectivity of MNV recovered from microplate wells containing medium and MB at different concentrations and exposed to a 12,500-lux red light source (light box) was analysed in RAW 264.7 cells. The cell culture limit of detection (LOD) was 1.8, and 2.8 log_10_ TCID_50_/mL for 100 µM MB and 1000 µM MB concentrations, respectively. All assays were performed as biological triplicates and technical duplicates (n=6). Means log_10_ TCID_50_/mL and standard deviations are represented. Values for positive controls (MB-untreated, but light box-exposed MNV) ranged between 5.69 and 6.19 log_10_ TCID_50_/mL.

### Murine norovirus inactivation on surgical masks via methylene blue photochemical treatment is concentration- and time-dependent

To establish optimal applied treatment conditions on the actual PPE items themselves, LB exposure time- and concentration-dependent virucidal kinetics of MB were investigated on SMs experimentally inoculated with MNV. 100 µM and 1000 µM methylene blue solution were applied to inoculated SMs which were subsequently subjected to 2 hours, 3 hours, or 4 hours of LB exposure. Limits of detections of both series of experiments were respectively lowered by one order of magnitude in these assays (0.8 instead of 1.8 and 1.8 instead of 2.8 log_10_ TCID_50_/mL for 100 µM and 1000 µM concentrations, respectively; effect attributed to a dilution of cytotoxic MB during elution), allowing determination of virus titre reductions of close to or more than four orders of magnitude for 100 µM concentrations at all three exposure times (3.96 (± 0.29), 4.10 (± 0.00), and 4.13 (± 0.00) log_10_ TCID_50_/mL) and over three orders of magnitude for 1000 µM concentrations (3.20 (± 0.00), 3.10 (± 0.00), 3.33 (± 0.14) log_10_ TCID_50_/mL) (Figure 4). At 1000 µM concentrations, MB solutions were macroscopically seen to undergo a (subjective) colour change from blue to violet (putatively indicative of aggregation) and to become viscous and agglomerate on SM surfaces, thus slowing the drying process.

**Figure 4.**
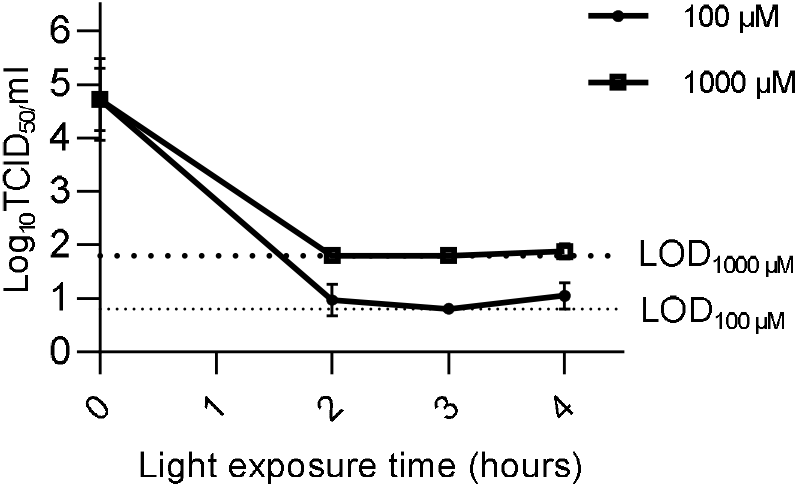
Evaluation of concentration- and time-dependent virucidal kinetics of methylene blue (MB) photochemical treatment on murine norovirus (MNV) – inoculated surgical masks. The infectivity of MNV recovered from surgical mask coupons treated with 100 µM or 1000 µM MB concentrations and subsequently exposed to a 12,500-lux red light source (light box) was analysed in RAW 264.7 cells. The cell culture limits of detection (LOD) were 0.8 and 1.8 log_10_ TCID_50_/mL for 100 µM and 1000 µM MB concentrations, respectively. All assays were performed as biological triplicates (n=3). Means log_10_ TCID_50_/mL and standard deviations are represented. Values for positive controls (MB-untreated, but light box-exposed MNV) ranged between 4.90 and 5.71 log_10_ TCID_50_/mL.

### Murine norovirus-inoculated surgical masks are reliably decontaminated via photochemical treatment involving 100 µM methylene blue coating of masks followed by 30 minutes of 12,500-lux red light exposure

To avoid issues associated with 1000 µM methylene blue concentrations (higher assay LOD, slow drying, putatively shifted absorption spectrum), further PPE-applied assays included only the 100 µM dye concentration. Based on the high reductions of infectious MNV titres of over three orders of magnitude from a 2-hour exposure onwards and in view of potentially achieving faster decontamination turn-around, shorter exposure times (90 minutes, 60 minutes, 30 minutes) were tested in a small preliminary assay (results not shown). Since high viral titre reductions were already observed following a 30-minute exposure in this preliminary assay, the 30-minute exposure was then tested and validated in four individual biological repeats (technical triplicates) (Figure 5). In addition, 15-minute LB exposures were assayed as biological and technical triplicates (Figure 6). While both the 30-minute- and the 15-minute protocol reduced mean infectious MNV titres by more than four orders of magnitude (4.71 (± 0.10) log_10_ TCID_50_/mL and 4.53 (± 0.23) log_10_ TCID_50_/mL, respectively), the shorter LB exposure led to higher inter-SM (and inter-coupon) variability (Figure 6) and did not consistently reduce virus titres to below the assay LOD.

**Figure 5.**
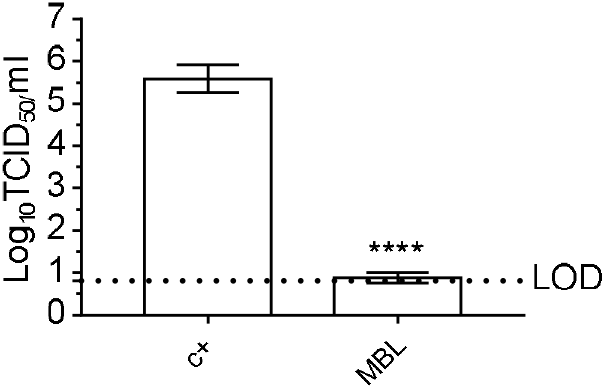
Evaluation of methylene blue (MB) photochemical treatment on murine norovirus (MNV) – inoculated surgical masks. The infectivity of MNV recovered from mask coupons treated with 100 µM MB and subsequently exposed to a 12,500-lux red light source (light box) for 30 minutes was analysed in RAW 264.7 cells. The cell culture limit of detection (LOD) was 0.8 log10 TCID_50_/mL. All assays were performed as biological quadruplicates and technical triplicates (n=12). Means log_10_ TCID_50_/mL and standard deviations are represented. Values for positive controls (MB-untreated, but light box-exposed MNV) ranged between 5.80 and 6.05 TCID_50_/mL. P-values were computed by using a two-sided independent sample t-test (where ****P<0.0001).

**Figure 6.**
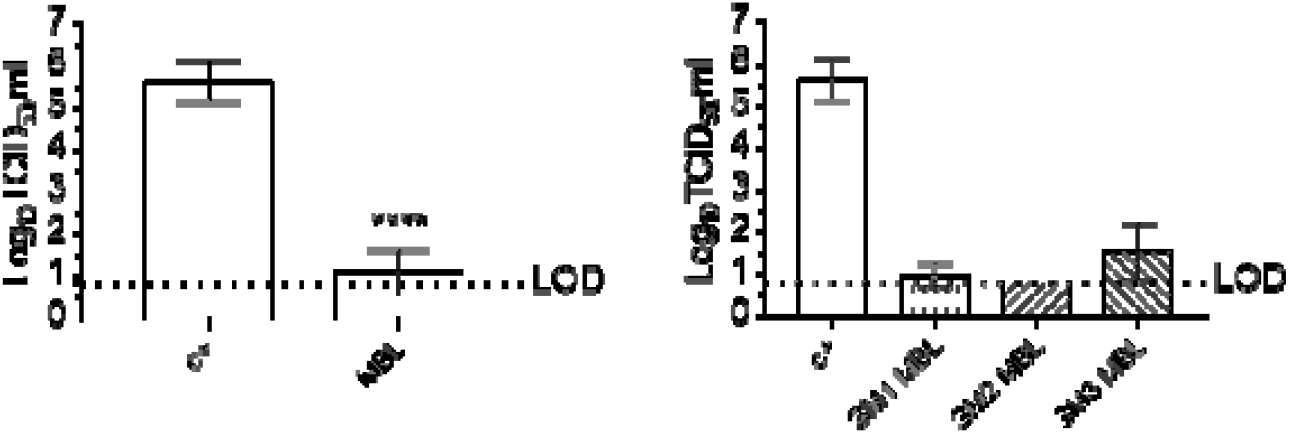
Evaluation of methylene blue (MB) photochemical treatment on murine norovirus (MNV) – inoculated surgical masks. The infectivity of MNV recovered from mask coupons treated with 100 µM MB and subsequently exposed for 15 minutes to a 12,500-lux red light source (light box) was analysed in RAW 264.7 cells. The cell culture limit of detection (LOD) was 0.8 log_10_ TCID_50_/mL. All assays were performed as biological and technical triplicates (n=9). The left panel represents a summary analysis of all obtained values. The right panel shows results for individual masks (n=3) and illustrates the varying decontamination efficacies of this protocol. Means log_10_ TCID_50_/mL and standard deviations are represented. Values for positive controls (MB-untreated, but light box-exposed MNV) ranged between 5.05 and 6.55 TCID_50_/mL. P-values were computed by using a two-sided independent sample t-test (where ****P<0.0001).

## DISCUSSION AND CONCLUSIONS

Supply issues at the beginning of the COVID-19 pandemic impressively illustrated that the world at large needs to be better positioned to deal quickly with prospective, potentially unknown, disease-causing agents and sanitary crises. Decontamination methods developed now, and perforce primarily targeting SARS-CoV-2, should thus already be future-proofed at this time by testing them against hardier pathogens. Here, we adapted an inexpensive and universally accessible photochemical decontamination protocol, recently developed against SARS-CoV-2 and other coronaviruses (DeMaND study) [15], to the treatment of norovirus-inoculated surgical masks.

To validate previously published MB photochemical treatment conditions [15] and to set a “baseline” for further development, the DeMaND MBL protocol involving 10 µM MB and 30-minutes of LB exposure was first investigated on either PRCV- or MNV-inoculated SMs. While this treatment reduced PRCV titres by over five orders of magnitude, it did not lead to significant inactivation of MNV.

Ten- and hundred-fold higher MB concentrations were subsequently tested in combination with longer LB exposures, first in a series of microplate-based assays and subsequently on MNV-inoculated SMs. In line with previous assays reporting oxygen-dependent laser inactivation of MNV (in solution) after long LB exposure times [24], inactivation of MNV proved both concentration- and time dependent in both matrices. In microplates, 100 µM MB concentrations markedly improved infectious titre reductions; 1000 µM MB solutions, however, did not further visibly enhance infectivity losses, this likely attributable to a saturation effect (the assay LOD and a minimum 4.80 (± 1.01) log_10_ TCID_50_/mL titre reduction were attained with 100 µM MB after 3 hours of LB exposure). Interestingly, MNV inactivation proved to be significantly more efficacious on SMs than in microplates and titre reductions of close to four orders of magnitude were already reached following 2 hours of LB exposure in initial SM assays.

The differences between MNV inactivation in microplates and SMs may be attributable to two not mutually exclusive effects. Thus, the LB-emitted light may not have been sufficiently powerful as to penetrate a highly concentrated 100 µM solution in deep microplate wells, whereas MB sprayed as a thin surface layer onto SMs was easily accessible to photons, presenting better opportunities for excitation and singlet oxygen production. Equally, since the concentration of oxygen in air is one order of magnitude greater than in water [25], a thin liquid-air interface (MB spray on SMs) could have positively impacted singlet oxygen production, whereas a larger-volume liquid phase (MB solution in microplate wells) may have had a negative impact.

Consolidated tests demonstrated that a MBL protocol involving 100 µM MB and a 30-minute LB exposure reliably reduced infectious MNV titres by more than four orders of magnitude. Shorter LB exposure led to higher inter-SM (and inter-coupon) variability and did not as consistently reduce MNV titres to below the assay LOD. A protocol involving shorter LB exposure times may be achievable via optimisation of MB dispersion (greater homogeneity via standardised nebuliser may improve virus inactivation); meanwhile, the 30-minute LB exposure in conjunction with simple MB application via spray bottle presents a low-cost and low-tech protocol and is recommended at this time for the inactivation of small non-enveloped viruses. The light box utilised in this study is simply constructed and uses commercially available horticultural LED lamps; further simplification of the method may be achieved by eliminating the need for light boxes entirely – the possibility of leveraging solar irradiation for MB activation is currently under investigation by other teams within the DeMaND consortium.

The precise mode of action of MB is yet to be to determined [16]; while the vastly different sensitivities between PRCV and MNV may implicate the viral envelope as being one of the targets of MBL treatment, varying densities of viral proteinaceous capsids as well as differences in viral genome size and corresponding susceptibility to nucleic acid strand breakage (at 7.4 kb MNV is roughly four times smaller than PRCV) may also play a role. Further studies are indicated to pinpoint the definitive virucidal effect(s) of photochemical decontamination.

Methylene blue photochemical treatment is easily adaptable to other SM or FFR types [15]. Owing to the variation in SM and FFR models (each FFR model has distinct filter materials and design characteristics which may result in variable outcomes of decontamination efficacy) the CDC recommends that the effectiveness of a decontamination be evaluated for specific FFRs in collaboration with the manufacturer, and if needed, a third-party laboratory. This holds true for inactivation of all contaminants and should ideally be performed for the norovirus-inactivating conditions tested in our study.

Methylene blue concentrations necessary for norovirus inactivation (the total dose of MB per SM is 0.24 mg), are ten-fold higher than those needed for coronavirus decontamination. Since there is considerable clinical experience to support that MB exhibits good safety [26,27] and MB concentrations used here were below those administered clinically, the use of this higher dose is unlikely to pose safety concerns. In addition, ultraviolet spectroscopy analyses testing the amount of MB that may leach off SMs and be inhaled by a wearer during the course of a ten-hour healthcare provider work shift, have shown that MB does not leach off SM (or other PPE) materials (unpublished data; ongoing project with the WHO). Briefly, a panel of SMs, FFRs, and a cloth community mask were subjected to a total of five 1000 µM MB treatment cycles (1000 µM MB solution; total volume: 35 – 40 mL MB applied). Treated and excised PPE coupons were then subjected to full-mask equivalent airflow rates of 120 L/minute with a total airflow of 43,200 L/coupon. With a level of detection of 0.004 mg/m^3^, no MB was observed within the spectroscopy parameters for all tested PPE items.

This is the first description of stable MB photochemical decontamination of SMs contaminated with an infectious norovirus. We describe successful validation of MB photochemical treatment for inactivation of small non-enveloped viruses that exceeds current FDA policy recommendations [5]. The highly resistant MNV surrogate supplements existing data regarding photochemical decontamination of SMs. It serves to future-proof this method against viral mask (and other PPE) contaminants, thus solidifying the position of MBL PPE treatment both to combat PPE shortages in austere or low resource environments and as an important tool in the global package of practical pandemic preparedness.

## Data Availability

All data produced in the present work are contained in the manuscript

## ACKNOWLEDGEMENTS

We thank May C. Chu (Department of Epidemiology, Colorado School of Public Health, Aurora, Colorado, USA) for her excellent management of the DeMaND consortium and her valuable help in setting up the DeMaND2 project. The authors express their sincere gratitude to Professor Hans Nauwynck (Laboratory of Virology, Faculty of Veterinary Medicine, Ghent University, Belgium) for the generous gift of both ST cells and PRCV strain 91V44. Within the framework of the Infection Prevention and Control (IPC) and World Health Emergencies (WHE) programmes of the World Health Organisation (WHO), we are grateful to Leandro Pecchia, Madison Moon, Brian Adams, Lisa Umphrey, Benedetta Allegranzi, and April Baller for their administrative advice and guidance throughout this project.

## Non-standard abbreviations

FFR: filtering facepiece respirator
SM: surgical mask
MNV: murine norovirus
SARS-CoV-2: severe acute respiratory syndrome coronavirus 2

## REFERENCES

[1] Strasser BJ, Schlich T. A history of the medical mask and the rise of throwaway culture. Lancet 2020;396:19–20. https://doi.org/10.1016/S0140-6736(20)31207-1.

[2] Ibn-Mohammed T, Mustapha KB, Godsell J, Adamu Z, Babatunde KA, Akintade DD, et al. A critical review of the impacts of COVID-19 on the global economy and ecosystems and opportunities for circular economy strategies. Resour Conserv Recycl 2021;164:105169. https://doi.org/10.1016/j.resconrec.2020.105169.

[3] World Health Organization (WHO). Rational use of personal protective equipment for coronavirus disease 2019 (COVID-19). Who 2020;2019:1–7.

[4] Implementing Filtering Facepiece Respirator (FFR) Reuse, Including Reuse after Decontamination, When There Are Known Shortages of N95 Respirators. Centers Dis Control Prev 2020. https://www.cdc.gov/coronavirus/2019-ncov/hcp/ppe-strategy/decontamination-reuse-respirators.html (accessed October 6, 2020).

[5] Center for Devices and Radiological Health. Enforcement Policy for Face Masks and Respirators During the Coronavirus Disease (COVID-19) Public Health Emergency (Revised) Guidance for Industry and Food and Drug Administration Staff 2020.

[6] Croke L. Preparing for the next infectious disease pandemic. AORN J 2020;112:P12–4. https://doi.org/10.1002/aorn.13188.

[7] Kramer A, Schwebke I, Kampf G. How long do nosocomial pathogens persist on inanimate surfacesã A systematic review. BMC Infect Dis 2006;6:1–8. https://doi.org/10.1186/1471-2334-6-130.

[8] Robilotti E, Deresinski S, Pinsky BA. Norovirus. Clin Microbiol Rev 2015;28:134–64. https://doi.org/10.1128/CMR.00075-14.

[9] Zonta W, Mauroy A, Farnir F, Thiry E. Comparative Virucidal Efficacy of Seven Disinfectants Against Murine Norovirus and Feline Calicivirus, Surrogates of Human Norovirus. Food Environ Virol 2015. https://doi.org/10.1007/s12560-015-9216-2.

[10] Ludwig-Begall LF, Mauroy A, Thiry E. Noroviruses—The State of the Art, Nearly Fifty Years after Their Initial Discovery. Viruses 2021;13:1541.

[11] Nims RW, Zhou SS. Intra-family differences in efficacy of inactivation of small, non-enveloped viruses. Biologicals 2016;44:456–62. https://doi.org/10.1016/j.biologicals.2016.05.005.

[12] Ludwig-Begall LF, Wielick C, Dams L, Nauwynck H, Demeuldre P-F, Napp A, et al. The use of germicidal ultraviolet light, vaporised hydrogen peroxide and dry heat todecontaminate face masks and filtering respirators contaminated with a SARS-CoV-2 surrogate virus. J Hosp Infect 2020. https://doi.org/10.1016/j.jhin.2020.08.025.

[13] Wielick C, Ludwig-Begall LF, Dams L, Razafimahefa RM, Demeuldre P-F, Napp A, et al. The use of germicidal ultraviolet light, vaporised hydrogen peroxide and dry heat to decontaminate face masks and filtering respirators contaminated with an infectious norovirus. Infect Prev Pract 2020;3:100111. https://doi.org/10.1016/j.infpip.2020.100111.

[14] Ludwig-Begall LF, Wielick C, Jolois O, Dams L, Razafimahefa RM, Nauwynck H, et al. “Don, doff, discard” to “don, doff, decontaminate” – FFR and mask integrity and inactivation of a SARS-CoV-2 surrogate and a norovirus following multiple vaporised hydrogen peroxide-, ultraviolet germicidal irradiation-, and dry heat decontaminations. PLoS One 2021.

[15] Lendvay TS, Chen J, Harcourt BH, Scholte FEM, Lin YL, Kilinc-Balci FS, et al. Addressing personal protective equipment (PPE) decontamination: Methylene blue and light inactivates SARS-COV-2 on N95 respirators and medical masks with maintenance of integrity and fit. Infect Control Hosp Epidemiol 2021;2019:1–10. https://doi.org/10.1017/ice.2021.230.

[16] Costa L, Faustino MAF, Neves MGPMS, Cunha Â, Almeida A. Photodynamic inactivation of mammalian viruses and bacteriophages. Viruses 2012;4:1034–74. https://doi.org/10.3390/v4071034.

[17] Eickmann M, Gravemann U, Handke W, Tolksdorf F, Reichenberg S, Müller TH, et al. Inactivation of Ebola virus and Middle East respiratory syndrome coronavirus in platelet concentrates and plasma by ultraviolet C light and methylene blue plus visible light, respectively. Transfusion 2018;58:2202–7. https://doi.org/10.1111/trf.14652.

[18] Seghatchian J, Walker WH, Reichenberg S. Updates on pathogen inactivation of plasma using Theraflex methylene blue system. Transfus Apher Sci 2008;38:271–80. https://doi.org/10.1016/j.transci.2008.04.004.

[19] Genina EA, Bashkatov AN, Chikina EE, Knyazev AB, Mareev O V., Tuchin V V. Methylene blue mediated laser therapy of maxillary sinusitis. Laser Phys 2006;16:1128–33. https://doi.org/10.1134/s1054660x06070188.

[20] McClurkin AW, Norman JO. Studies on transmissible gastroenteritis of swine. II. Selected characteristics of a cytopathogenic virus common to five isolates from transmissible gastroenteritis. Can J Comp Med Vet Sci 1966;30:190–8.

[21] Cox E, Hooyberghs J, Pensaert MB. Sites of replication of a porcine respiratory coronavirus related to transmissible gastroenteritis virus. Res Vet Sci Sci 1990;48:165–169.

[22] Reed, L.J.; Muench H. A simple method of estimating fifty percent endpoints. Am J Hyg 1938;27.

[23] Lendvay TS, Chen J, Harcourt BH, Scholte FEM, Kilinc-Balci FS, Lin YL, et al. Addressing Personal Protective Equipment (PPE) DecontaminationL: Methylene Blue and Light Inactivates SARS-CoV-2 on N95 Respirators and Masks with Maintenance of Integrity and Fit. MedRxiv 2020.

[24] Kingsley D, Kuis R, Perez R, Basaldua I, Burkins P, Marcano A, et al. Oxygen-dependent laser inactivation of murine norovirus using visible light lasers. Virol J 2018;15:1–8. https://doi.org/10.1186/s12985-018-1019-2.

[25] Fondriest Environmental Inc. Dissolved Oxygen -Fundamentals of Environmental Measurements 2013. https://www.fondriest.com/environmental-measurements/parameters/water-quality/dissolved-oxygen.

[26] Seghatchian J, Struff WG, Reichenberg S. Main properties of the THERAFLEX MB-plasma system for pathogen reduction. Transfus Med Hemotherapy 2011;38:55–64. https://doi.org/10.1159/000323786.

[27] Rengelshausen J, Burhenne J, Fröhlich M, Tayrouz Y, Singh SK, Riedel KD, et al. Pharmacokinetic interaction of chloroquine and methylene blue combination against malaria. Eur J Clin Pharmacol 2004;60:709–15. https://doi.org/10.1007/s00228-004-0818-0.

